# Effectiveness of the offer of the *Smoke Free* smartphone application compared with no intervention for smoking cessation: a pragmatic randomised controlled trial

**DOI:** 10.1101/2023.01.12.23284463

**Authors:** Sarah E. Jackson, Dimitra Kale, Emma Beard, Olga Perski, Robert West, Jamie Brown

## Abstract

**Aim:** To evaluate the effectiveness of the offer of *Smoke Free* – an evidence-informed, widely used app – for smoking cessation versus no support.

**Design:** Two-arm individually randomised controlled effectiveness trial.

**Setting:** Online with no restrictions on location.

**Participants:** 3,143 adult smokers (74.7% female; mean[SD] age 49.0 [11.5] years) motivated to make a quit attempt in the next month, recruited between August-2020 and April-2021.

**Interventions and comparators:** Offer of the *Smoke Free* app plus follow-up (intervention arm) versus no intervention plus follow-up (comparator arm). Both groups were shown a brief message at the end of the baseline questionnaire encouraging them to make a quit attempt.

**Main outcome measures:** The primary outcome was self-reported 6-month continuous abstinence assessed 7 months after randomisation. Secondary outcomes included quit attempts in the first month post-randomisation, 3-month continuous abstinence assessed at 4 months, and 6-month continuous abstinence at 7 months among those who made a quit attempt. The primary analysis was performed on an intention-to-treat basis, with missing-equals-smoking imputation. Sensitivity analyses included i) restricting the intervention group to those who took up the offer of the app, ii) using complete cases, and iii) using multiple imputation.

**Results:** The effective follow-up rate for 7 months was 41.9%. The primary analysis showed no evidence of a benefit of the intervention on rates of 6-month continuous abstinence (intervention 6.8% vs. comparator 7.0%; RR=0.97, 95%CI=0.75-1.26). Analyses on all secondary outcomes also showed no evidence of a benefit. Similar results were observed on complete cases and using multiple imputation. When the intervention group was restricted to those who took up the offer of the app (n=395, 25.3%), participants in the intervention group were 80% more likely to report 6-month continuous abstinence (12.7% vs. 7.0%; RR=1.80, 95%CI=1.30-2.45). Equivalent subgroup analyses produced similar results on the secondary outcomes. These differences persisted after adjustment for key baseline characteristics.

**Conclusions:** Among motivated smokers provided with very brief advice to quit, offer of the *Smoke Free* app did not have a detectable benefit for cessation compared with follow-up only. However, the app increased quit rates when smokers randomised to receive the app downloaded it.

## Introduction

Tobacco remains one of the leading causes of disease and preventable death globally, killing in excess of 8 million people each year.^1^ A range of evidence-based support options are available to help smokers quit,^2–4^ but barriers including a lack of time and (in countries where free support is unavailable) inability to pay mean many smokers do not access this support.^5–7^ The Covid-19 pandemic has brought this issue into sharper focus: access to face-to-face stop smoking services was suspended or altered during periods of social restriction^8,9^ and the rate of unaided quit attempts increased.^10^ Digital technologies offer potential for low-cost, scalable delivery of interventions to promote smoking cessation. More smokers have turned to remote options for cessation support since the pandemic began, including smartphone applications (apps),^10^ but evidence on the effectiveness of existing apps is lacking. This study aimed to assess the effectiveness of a widely used app for smoking cessation compared with no support.

Digital support for smoking cessation has the potential to contribute to meaningful reductions in smoking prevalence in countries around the world. In particular, the past decade has seen a surge in smartphone apps offering support for smokers who want to quit.^11^ Combined with increasing levels of smartphone ownership (currently estimated at 3.8 billion worldwide;^12^ 74% of the adult population^13^), these apps can reach large numbers of smokers. However, the potential of apps for promoting cessation is not yet being realised. A 2019 Cochrane review of mobile phone interventions for smoking cessation^14^ identified just five studies (total n=3,079) that compared a smoking cessation smartphone app with lower-intensity smoking cessation support (either a lower-intensity app or non-app minimal support). The pooled data provided no evidence that smartphone apps increased the likelihood of smoking cessation (relative risk [RR]=1.00, 95% confidence interval [CI]=0.66-1.52), but the evidence was judged to be of very low certainty which limits confidence in the effect estimate. The authors called for more large-scale randomised controlled trials (RCTs) to establish whether smartphone app interventions are effective for smoking cessation. In 2020, a large RCT^11^ (n=2,415) tested the efficacy of a smoking cessation app based on acceptance and commitment therapy (ACT) compared with a simpler app informed by United States clinical practice guidelines. Self-reported quit rates at 12 months were higher among participants randomised to use the ACT-based app (28.2% vs. 21.1%, odds ratio [OR]=1.49, 95%CI=1.22-1.83). To our knowledge, no published RCTs have compared apps designed to provide ongoing support with unaided quitting.

With over 6 million downloads to date and 70,000 new users each month, *Smoke Free* (smokefreeapp.com) is one of the world’s most widely used smoking cessation apps. The app is evidence-informed and available for iOS and Android OS. In a large exploratory RCT (n=28,112) conducted between 2013 and 2015, the full version of the app increased 3-month self-reported continuous abstinence rates compared with a reduced version (OR 1.90, 95%CI=1.53-2.37).^15^ However, this result was limited by relatively short-term outcomes and very low follow-up rates (8.5% in the intervention condition and 6.5% in the control condition) with no active attempts to recontact participants. *Smoke Free* therefore constitutes a useful test bed for assessing the effectiveness of a smartphone app for smoking cessation versus unaided quitting.

This paper describes the results of the *App for Smoking ceSsation Evaluation Trial (ASSET)*,^16^ a two-arm RCT designed to evaluate the effectiveness of the offer of the *Smoke Free* app in increasing rates of tobacco smoking cessation compared with follow-up only. We adopted a pragmatic design in order to provide information on the usefulness of this app in real-world settings. Given that, in an effectiveness trial of this nature, not all participants who are offered the intervention will take it up, we were therefore testing the offer rather than actual use of the app. The primary research question was:

1. In English-speaking adult smokers willing to quit in the next 4 weeks who are recruited online or have previously used the *Smoke Free* app and agreed to be followed-up (population) and in an unrestricted online location (setting), how effective is an offer to use the app plus follow-up (intervention) compared with no offer of the app and follow-up only (comparator) in promoting self-reported smoking cessation for at least 6 months, assessed 7 months after enrolment (outcome 1)? We also addressed a number of secondary research questions:
2. In the population, setting and intervention versus comparator for RQ1, what is the effect in promoting at least one quit attempt in the 1 month following enrolment in the study (outcome 2)?
3. In the population, setting and intervention versus comparator for RQ1, what is the effect in promoting smoking cessation for at least 3 months, assessed 4 months after enrolment (outcome 3)?
4. In the setting, intervention versus comparator and outcome for RQ1, what is the effect in those who make at least one quit attempt in the 1 month following enrolment in the study (population 2)?
5. In the population, setting and intervention versus comparator for RQ1, what is the effect in promoting downloading or using the *Smoke Free* app at least once, assessed 7 months after enrolment (outcome 4)?
6. Are the answers to RQs 1–5 different in smokers who are:
  a. male versus female (population moderator 1);
  b. more versus less addicted to cigarettes (population moderator 2);
  c. aged 18–34 versus ≥35y (population moderator 3);
  d. with versus without post-16 educational qualifications (population moderator 4);
  e. from poorer versus richer financial situations (population moderator 5); and
  f. have previous versus no prior experience with the app (population moderator 6)?

## Method

A summary timeline of trial procedures is shown in **Supplementary File 1**.

### Design

ASSET was a two-arm individually randomised controlled trial. The study protocol^16^ and analysis plan were pre-registered (ISRCTN85785540; https://osf.io/umec4). An independent Trial Steering Committee provided overall supervision of the trial.

### Setting

The study was conducted online using the Qualtrics survey platform, with no restriction on participants’ location.

### Participants

Participant inclusion criteria were: current cigarette smoker; aged ≥18y; English speaker; owns a smartphone; provided a valid email address not previously used by another participant; interested in making a quit attempt within the next month; willing to be followed up by email and complete online questionnaires 1, 4, and 7 months after enrolling in the study; able to provide consent.

Eligibility was assessed via screening questions embedded at the start of the baseline study questionnaire on Qualtrics. Those who did not meet the inclusion criteria were informed that they were not eligible to participate and directed to the NHS Smokefree website (https://www.nhs.uk/smokefree) for resources to help with quitting smoking.

Participants were not provided with any financial compensation for taking part in the study, but we offered to donate £10 (US$12) to a cancer charity on behalf of each participant who responded to the final follow-up.

### Sample size

The intended sample size was decided a priori based on achieving 90% power to detect a RR of ≥1.5 with an alpha of *p*<0.05 one-tailed and a quit rate of 6.0% in the comparator group. This led to a target sample size of 3,116; 1,558 in each group.

We note that we amended our original power calculation that had a larger target sample size as reported in the published protocol.^16^ Details of these amendments and the rationale underlying them are available on Open Science Framework (https://osf.io/umec4). The final sample size target was approved by the Trial Steering Committee on 6 January 2021 and registered on ISRCTN in January 2021 after we had randomised 2,798 participants and before any of the primary outcome data (6-month continuous abstinence assessed at 7 months) were collected.

### Ethical approval

Ethical approval for the trial was obtained from the UCL Research Ethics Committee (reference: CEHP/2020/579). All participants were provided with a summary of the study and their right to withdraw on the landing page of the baseline survey on Qualtrics. They provided informed consent by selecting ‘Yes: I confirm I have read the information about the study and wish to participate’.

### Recruitment

Recruitment took place between August 2020 and April 2021. Participants were recruited via advertisements on social media (Facebook and Twitter; **Supplementary File 1**) and a mailing list of smokers who had previously signed up to the *Smoke Free* app and had agreed to be contacted. We emailed people on the mailing list who signed up to the app >6 months previously with an invitation to participate in the study. Response to these emails was low (∼5 in 1000). The majority of our participants were recruited via Facebook adverts (at an average cost of £3/participant) which linked to the Qualtrics baseline questionnaire (with embedded consenting procedure and screening questions).

### Randomisation

People who consented to participate and met the eligibility criteria were randomised after completing the baseline questionnaire to either the intervention or comparator condition. Randomisation was 1:1 at the individual level with no restriction (i.e., no blocking) and was automated within Qualtrics, such that each participant was shown at random either the intervention message including offer of the *Smoke Free* app or the comparator message after the final questionnaire question.

All investigators were blinded to participants’ treatment allocation until all data had been collected. The data were analysed blind by the trial statistician (EB).

### Interventions

#### Comparator

After consenting and completing the baseline questionnaire, participants in the comparator condition were shown a final screen with a brief message encouraging them to make a quit attempt within the next 4 weeks and reminding them of the importance of responding to follow-up requests designed to track their progress (**Supplementary File 1**). This same message was also emailed to them immediately afterwards.

#### Intervention: Smoke Free app

Participants in the intervention condition received the same advice as those in the comparator condition plus offer of the full version of the *Smoke Free* app free of charge, encouragement to use the app, and a link to download it. This same message and information on how to access the app was also emailed to them immediately afterwards.

The *Smoke Free* app is based on behaviour change techniques that would be expected from theory^15^ and evidence with face-to-face support^17,18^ to aid smoking cessation. It guides smokers through the first three months of their quit attempt by helping them maintain their resolve by setting a clear goal, monitoring their progress towards that goal and becoming aware of benefits of being smoke-free achieved to date. The app has several components:

1. A calculator which tracks the total amount of money not spent on buying cigarettes and the number of cigarettes not smoked (‘Dashboard’);
2. A calendar which tracks the amount of time elapsed since cessation (‘Dashboard’);
3. A scoreboard which awards virtual ‘badges’ to users for not smoking (‘Badges’);
4. Progress indicators which inform users of the health improvements made since the start of their quit attempt (e.g. pulse rate, oxygen levels, carbon monoxide levels; ‘Dashboard’);
5. A diary which tracks the frequency, strength, location and triggers of cravings to smoke (‘Diary’);
6. A graph which displays the frequency, location, strength and triggers of cravings to smoke (‘Cravings’);
7. Daily missions which are assigned from the start of a user’s quit date for one calendar month (‘Missions’);
8. A chatbot which delivers evidence-based guidance about quitting smoking via a conversational interface which resembles text messaging;
9. 24/7 access to National Centre for Smoking Cessation and Training (NCSCT)-trained advisors;
10. Advisor-led stop smoking clinics held in-app four times a day.

Behaviour change techniques included in the app are summarised in the trial protocol.^16^

### Follow-up data collection

Follow-up data were collected between September 2020 and December 2021, via online questionnaires 1, 4, and 7 months after study enrolment. For the 1- and 4-month follow-ups, participants were invited to complete the survey via email. This was automated within Qualtrics. For the final (7-month) follow-up, in order to boost response rates for our primary outcome, we contacted participants up to six times over two weeks. First, they were invited via email within Qualtrics. Next, a further email invitation was sent from one of the research team’s personal email address (in an effort to reduce the ‘spam’ rating of the email). Then participants who provided their phone number were contacted via SMS (between March and August 2021) or telephone call (between August and December 2021; a change implemented in an effort to boost the response rate) and asked to respond ‘yes’ or ‘no’ to the key outcome assessment. Finally, up to three further emails were sent asking the same question as the SMS, prompting participants for a direct ‘yes’ or ‘no’ response via email. Participants who responded to any of the invitations/reminders were not contacted further.

### Measures

#### Participant baseline characteristics

The baseline questionnaire assessed the following: email address, mobile phone number (optional), smartphone ownership, motivation to quit in the next month (Motivation To Stop Scale^19^), willingness to complete online questionnaires after 1, 4 and 7 months, age (18-34/35-64/≥65 years), gender (male/female), education (any/no post-16 qualifications), financial status (live comfortably/meet needs with a little left/just meet basic expenses/don’t meet basic expenses),^20^ country of residence, first language (English/other), number of cigarettes smoked per day, level of cigarette addiction (first cigarette within 5 minutes/6-30 minutes/31-60 minutes/>60 minutes of waking), history of serious quit attempts (never/yes – not in the past year/yes – in the past year), and past and current use of support for smoking cessation (prescription nicotine replacement therapy (NRT)/NRT bought over-the-counter/varenicline/bupropion/face-to-face behavioural support/telephone support/written self-help materials/websites/apps). In an exploratory addition to outcome assessment, given evidence that heart rate declines substantially when smokers stop,^21^ participants who had a heart rate monitoring device (e.g., FitBit/Apple watch) were asked to report their average resting heart rate as indicated by their app.

#### Outcomes

The primary outcome was the percentage of participants reporting not having smoked for 6 months at the 7-month follow-up. This was assessed in the online questionnaire with the question: *‘Have you smoked any cigarettes in the past 6 months?’* with response options *‘none at all’, ‘between 1 and 5’*, and *‘more than 5’*. In line with the Russell Standard for self-report of smoking abstinence,^22^ the former two responses were collapsed for analysis, with data coded 1 for respondents reporting smoking no more than 5 cigarettes in the past 6 months and 0 for those reporting smoking more than 5 cigarettes. Where participants did not respond to the invitations to complete the questionnaire, the question was simplified to *‘Have you smoked more than 5 cigarettes in the past 6 months?’* and participants were asked to reply *‘yes’* or *‘no’* via SMS, telephone, or email. On the basis of the intention-to-treat principle, Bthose who did not respond to follow-up attempts were retained in the analyses and classified as continuing smokers.^22^

Secondary outcomes were the percentage of participants reporting:

1. Quit attempts at 1-month follow-up, defined as having made a serious quit attempt in the last 4 weeks (assessed with the question: *‘Have you made a serious attempt to quit smoking in the last 4 weeks? Please include any attempt that you are currently making [yes=1/no=0]’;*
2. Smoking cessation for at least 3 months at the 4-month follow-up (assessed with the question: *‘Have you smoked a single puff on a cigarette in the past 3 months? [yes=0/no=1]’);*
3. Smoking cessation for at least 6 months at the 7-month follow-up in those who made a quit attempt (assessed with the question: *‘Have you smoked any cigarettes in the past 6 months? [none at all=1/between 1 and 5=1/more than 5=0]’);*
4. App use, defined as downloading or using the *Smoke Free* app at least once at any point during the study period (assessed at the 7-month follow-up with the question: *‘In the last 7 months, have you downloaded or used the Smoke Free app (pictured) at least once? [yes=1/no=0]’)*.

### Statistical analyses

We followed our pre-registered analysis plan^16^, with two amendments registered on Open Science Framework prior to running the analysis (https://osf.io/umec4/) and two unplanned sensitivity analyses after running the analyses. Details of amendments to the analysis plan are summarised in **Supplementary File 2**.

All variables were collected primarily online and entered automatically into a Qualtrics database. From this database, a user-specified Excel file was downloaded, subjected to basic processing and re-coding, and integrated with responses provided by text messages. On completion, data were analysed blind to intervention allocation using R Studio v.4.2.1.

#### Primary analyses

Our primary analyses used an intention-to-treat approach, with missing-equals-smoking imputation.^22^ We used log-binomial regression to calculate the RR and 95%CI of each primary and secondary outcome in the intervention group versus the comparator group.

#### Moderation analyses

For each primary and secondary endpoint, we ran a series of log-binomial regression models in which we added two-way interactions between group (intervention/comparator) and gender (male/female), cigarette addiction (first cigarette within 5 minutes/6-30 minutes/31-60 minutes/>60 minutes after waking), age (<35/≥35), education (post-16 qualifications: yes/no), financial situation (live comfortably/meet needs with a little left/just meet basic expenses/don’t meet basic expenses), and previous experience with using a smoking cessation app (previous experience: yes/no – based on self-reported ever use of cessation aids at baseline). Results are reported in **Supplementary File 3**.

#### Sensitivity analyses

We repeated our primary and secondary analyses (i) restricting the intervention group to participants who took up the offer of full free access to the *Smoke Free* app, which was self-reported or verified by matching the email address used to login to the app to the one provided in the baseline questionnaire (with and without adjustment for key baseline characteristics); (ii) restricting both groups to participants who were successfully followed up; and (iii) using multiple imputation to impute missing outcomes data. In an exploratory analysis, we also repeated the analyses defining successful quits as self-reported abstinence plus a reduction in mean resting heart rate of ≥5 beats per minute, based on the lower 95% CI for the difference in resting heart rate between people smoking as usual and not smoking in a previous study.^23^ Finally, we reanalysed our primary outcome assuming different rates of abstinence in those not followed-up.

In order to aid interpretation of the strength of evidence for associations, we calculated Bayes factors (BFs)^24^ to differentiate between evidence for an effect, no effect, and data insensitivity. We used a half-normal distribution, the mode at 0 (no effect), and the standard deviation equal to the expected effect size used in the sample size calculation (RR=1.5). BFs ≥3 can be interpreted as evidence for the alternative hypothesis, ≤1/3 as evidence for the null hypothesis, and BFs between 1/3 and 3 suggest the data are insensitive to distinguish the alternative hypothesis from the null.^25,26^

## Results

A total of 3,143 eligible participants were recruited, completed the baseline assessment, and were randomised to the intervention or comparator condition. **Figure 1** shows the numbers allocated to each group and followed-up and **Table 1** summarises baseline socio-demographic and smoking characteristics. Distribution of socio-demographic and smoking characteristics were similar across groups, consistent with successful randomisation.

**Figure 1.**
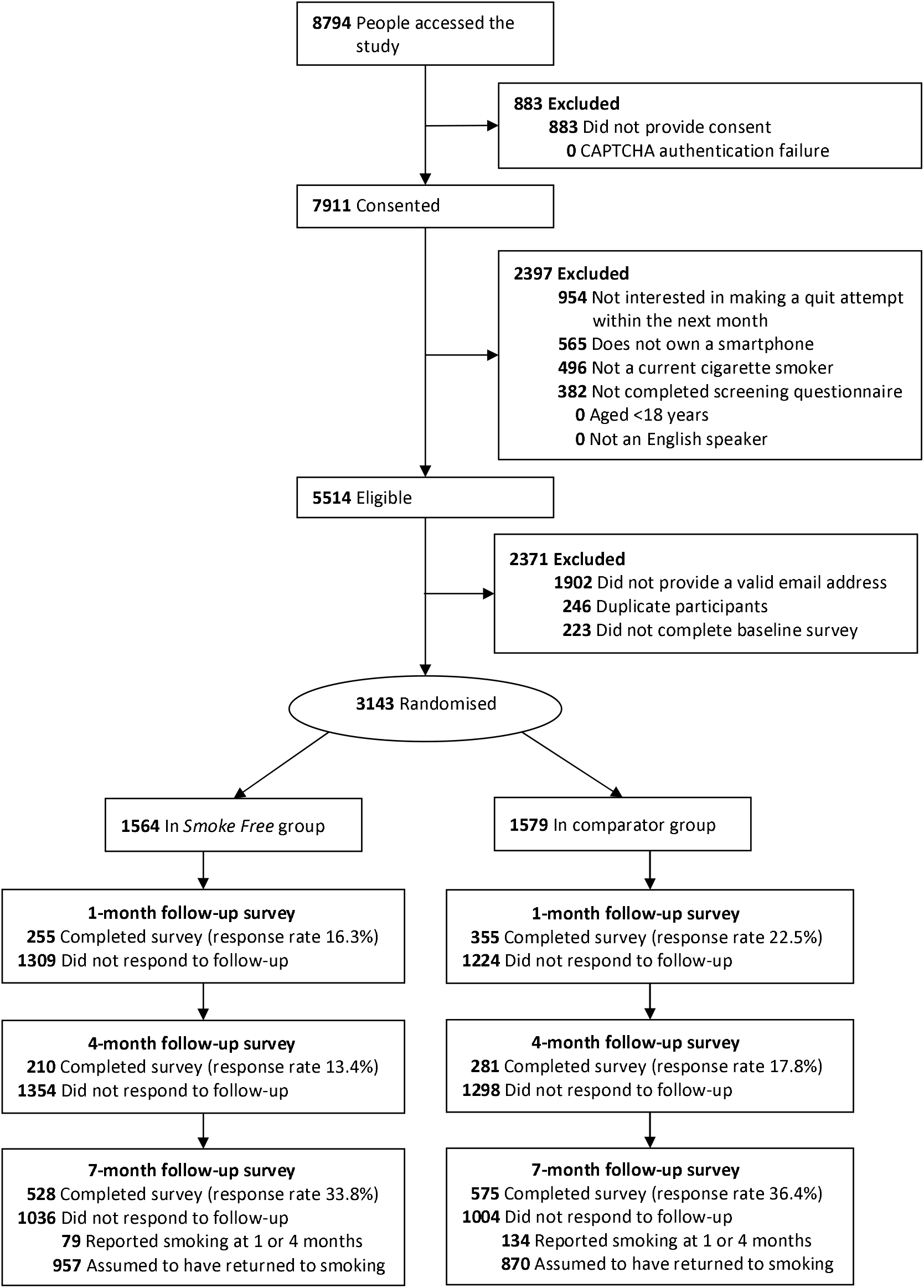
CONSORT flowchart.

**Table 1.** Baseline characteristics of participants randomised to each condition

|  | Total<br>(n=3143) |  | Comparator<br>(n=1579) |  | Offered *Smoke*<br>*Free app* (n=1564) |  |
| --- | --- | --- | --- | --- | --- | --- |
|  | % | n | % | n | % | n |
| Age (years) |  |  |  |  |  |  |
| 18-34 | 11.5 | 361 | 11.4 | 180 | 11.6 | 181 |
| 35-64 | 81.5 | 2562 | 82.0 | 1294 | 81.1 | 1268 |
| 65+ | 7.0 | 220 | 6.7 | 105 | 7.4 | 115 |
| Gender |  |  |  |  |  |  |
| Male | 25.0 | 785 | 24.2 | 382 | 25.8 | 403 |
| Female | 74.7 | 2349 | 75.5 | 1192 | 74.0 | 1157 |
| Other | 0.3 | 9 | 0.3 | 5 | 0.3 | 4 |
| Post-16 qualifications | 90.5 | 2845 | 90.4 | 1427 | 90.7 | 1418 |
| Financial status |  |  |  |  |  |  |
| Live comfortably | 5.6 | 176 | 5.9 | 93 | 5.3 | 83 |
| Meet needs with a little left | 32.8 | 1031 | 33.5 | 529 | 32.1 | 502 |
| Just meet basic expenses | 41.6 | 1307 | 39.7 | 627 | 43.5 | 680 |
| Don't meet basic expenses | 20.0 | 629 | 20.9 | 330 | 19.1 | 299 |
| Country of residence |  |  |  |  |  |  |
| UK | 45.4 | 1426 | 45.8 | 723 | 45.0 | 703 |
| USA | 33.9 | 1065 | 34.3 | 542 | 33.4 | 523 |
| Canada | 8.6 | 270 | 8.0 | 127 | 9.1 | 143 |
| Ireland | 6.4 | 201 | 5.8 | 91 | 7.0 | 110 |
| Australia | 3.0 | 95 | 3.0 | 48 | 3.0 | 47 |
| Other | 2.7 | 86 | 3.0 | 48 | 2.4 | 38 |
| English as first language | 95.4 | 2997 | 95.8 | 1513 | 94.9 | 1484 |
| Time to first cigarette |  |  |  |  |  |  |
| ≤5 minutes | 45.3 | 1424 | 46.0 | 726 | 44.6 | 698 |
| 6-30 minutes | 39.6 | 1246 | 40.2 | 635 | 39.1 | 611 |
| 31-60 minutes | 8.6 | 269 | 7.8 | 123 | 9.3 | 146 |
| >60 minutes | 6.5 | 204 | 6.0 | 95 | 7.0 | 109 |
| History of serious quit attempts |  |  |  |  |  |  |
| Never | 7.1 | 223 | 7.3 | 115 | 6.9 | 108 |
| Yes – not in the past year | 58.9 | 1852 | 59.6 | 941 | 58.3 | 911 |
| Yes – in the past year | 34.0 | 1068 | 33.1 | 523 | 34.9 | 545 |

**Table 1.** (continued)
|  | Total<br>(n=3143) |  | Comparator<br>(n=1579) |  | Offered *Smoke Free* app (n=1564) |  |
| --- | --- | --- | --- | --- | --- | --- |
|  | % | n | % | n | % | n |
| **Past use of cessation support** |  |  |  |  |  |  |
| Prescription NRT | 52.4 | 1648 | 53.1 | 839 | 51.7 | 809 |
| NRT bought over the counter | 29.6 | 930 | 30.0 | 474 | 29.2 | 456 |
| Varenicline | 16.5 | 518 | 15.6 | 246 | 17.4 | 272 |
| Bupropion | 14.6 | 460 | 14.1 | 222 | 15.2 | 238 |
| Face-to-face behavioural support | 8.2 | 257 | 8.7 | 138 | 7.6 | 119 |
| Telephone support | 6.2 | 195 | 5.9 | 93 | 6.5 | 102 |
| Written self-help materials | 23.9 | 751 | 24.3 | 384 | 23.5 | 367 |
| Websites | 10.6 | 333 | 11.0 | 173 | 10.2 | 160 |
| Apps | 48.7 | 1531 | 47.7 | 753 | 49.7 | 778 |
| E-cigarette or other vaping device | 16.4 | 516 | 17.0 | 269 | 15.8 | 247 |
| Other | 3.3 | 102 | 2.7 | 43 | 3.8 | 59 |
| None of the above | 12.8 | 402 | 13.1 | 206 | 12.5 | 196 |
| **Current use of cessation support** |  |  |  |  |  |  |
| Prescription NRT | 2.5 | 78 | 2.5 | 39 | 2.5 | 39 |
| NRT bought over the counter | 9.7 | 304 | 9.1 | 143 | 10.3 | 161 |
| Varenicline | 1.9 | 58 | 2.0 | 31 | 1.7 | 27 |
| Bupropion | 0.7 | 21 | 0.7 | 11 | 0.6 | 10 |
| Face-to-face behavioural support | 0.2 | 7 | 0.1 | 2 | 0.3 | 5 |
| Telephone support | 0.8 | 25 | 0.4 | 7 | 1.2 | 18 |
| Written self-help materials | 1.8 | 57 | 1.8 | 28 | 1.9 | 29 |
| Websites | 2.7 | 85 | 2.4 | 38 | 3.0 | 47 |
| Apps | 3.9 | 123 | 4.0 | 63 | 3.8 | 60 |
| E-cigarette or other vaping device | 12.2 | 382 | 12.3 | 194 | 12.0 | 188 |
| Other | 0.9 | 28 | 0.8 | 12 | 1.0 | 16 |
| None of the above | 69.8 | 2195 | 70.6 | 1114 | 69.1 | 1081 |
|  | Mean | SD | Mean | SD | Mean | SD |
| Age (years) | 49.0 | 11.5 | 48.9 | 11.4 | 49.2 | 11.6 |
| Cigarettes per day | 18.1 | 9.4 | 18.2 | 10.0 | 18.0 | 8.7 |
| Resting heart rate* | 75.2 | 18.2 | 75.4 | 18.7 | 74.9 | 17.6 |

NRT, nicotine replacement therapy. SD, standard deviation.
\* If participants had a heart monitoring device (e.g. Fitbit, Apple watch); this was not a required field (missing overall 72.73%, n=2286; *Smoke Free* 71.82% n=1134; comparator 73.66%, n=1152).

Response rates to the 1-month, 4-month and 7-month follow-ups were 19.4%, 15.6%, and 35.1% respectively; 16.3%, 13.4%, and 33.8% in the *Smoke Free* group, and 22.5%, 17.8%, and 36.4% in the comparator group (**Figure 1**). Of the 2040 participants who did not respond directly to the final follow-up, 213 (10.4%) individuals reported smoking in a previous follow-up assessment, which would have classified them as smokers by our continuous abstinence primary outcome, meaning the effective follow-up rate for the primary outcome was 41.9% (2040 – 213 = 1827; 3143 – 1827 = 1316; 1316/3143); 38.8% in the *Smoke Free* group and 44.9% in the comparator group.

### Primary outcome

**Table 2** shows results for the primary outcome of 6-month continuous abstinence from smoking, assessed at 7-month follow-up, using an intention-to-treat approach with missing-equals-smoking imputation. Overall, 6.9% of participants reported 6-month continuous abstinence. The rate of smoking cessation was similar between participants in the *Smoke Free* and comparator groups (6.8% in the intervention group vs. 7.0% in the comparator group). The Bayes factor favoured no effect. Moderation analyses showed no significant difference in treatment effect by any characteristic (**Supplementary File 3**).

**Table 2.**
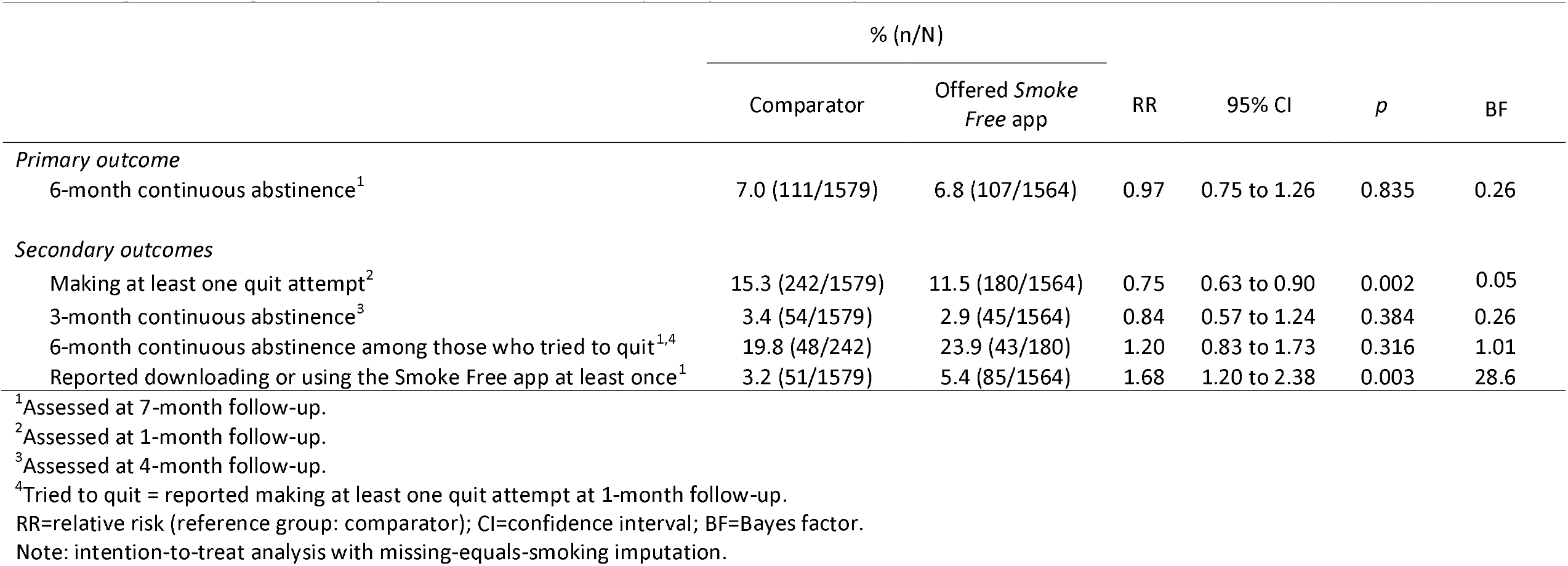
Log-binomial regression analyses of treatment effect on primary and secondary outcomes

|  | % (n/N) |  | RR | 95% CI | *p* | BF |
| --- | --- | --- | --- | --- | --- | --- |
|  | Comparator | Offered *Smoke Free* app |  |  |  |  |
| *Primary outcome* |  |  |  |  |  |  |
| 6-month continuous abstinence<sup>1</sup> | 7.0 (111/1579) | 6.8 (107/1564) | 0.97 | 0.75 to 1.26 | 0.835 | 0.26 |
| *Secondary outcomes* |  |  |  |  |  |  |
| Making at least one quit attempt<sup>2</sup> | 15.3 (242/1579) | 11.5 (180/1564) | 0.75 | 0.63 to 0.90 | 0.002 | 0.05 |
| 3-month continuous abstinence<sup>3</sup> | 3.4 (54/1579) | 2.9 (45/1564) | 0.84 | 0.57 to 1.24 | 0.384 | 0.26 |
| 6-month continuous abstinence among those who tried to quit<sup>1,4</sup> | 19.8 (48/242) | 23.9 (43/180) | 1.20 | 0.83 to 1.73 | 0.316 | 1.01 |
| Reported downloading or using the *Smoke Free* app at least once<sup>1</sup> | 3.2 (51/1579) | 5.4 (85/1564) | 1.68 | 1.20 to 2.38 | 0.003 | 28.6 |

<sup>1</sup>Assessed at 7-month follow-up.<sup>2</sup>Assessed at 1-month follow-up.<sup>3</sup>Assessed at 4-month follow-up.<sup>4</sup>Tried to quit = reported making at least one quit attempt at 1-month follow-up.
RR=relative risk (reference group: comparator); CI=confidence interval; BF=Bayes factor.
Note: intention-to-treat analysis with missing-equals-smoking imputation.

### Secondary outcomes

**Table 2** also summarises results relating to the secondary outcomes. Participants in the *Smoke Free* group had a 25% lower risk of reporting a quit attempt compared with those in the comparator group. There was no statistically significant difference in 3-month continuous abstinence rates between groups or 6-month continuous abstinence among those who tried to quit (Bayes factors for these outcomes favoured no effect). Those in the *Smoke Free* group were 68% more likely to report having downloaded or used the *Smoke Free* app at least once during the study period than those in the comparator group, although self-reported rates were very low in both groups (5.4% in the intervention group vs. 3.2% in the comparator group).

### Sensitivity analyses

**Table 3** shows results for primary and secondary outcomes restricting the intervention group to those who took up the offer of the *Smoke Free* app. Despite just 85 participants (5.4%) in the intervention group recalling, in the 7-month follow-up survey, having downloaded or used the *Smoke Free* app at least once, matching email address logins were verified for 355 participants (22.7%) in the intervention group – indicating that they had (at least briefly) taken up the offer of the app after the baseline survey. This discrepancy is likely to be largely due to loss to follow-up; the majority of participants either did not respond to the 7-month follow-up survey or only provided data on the primary outcome. Combined, a total of 395 participants (25.3%) in the intervention group either self-reported or had verified app use. **Supplementary File 4** compares the baseline characteristics of these intervention participants who took up the offer of the app with those of the comparator group. The 6-month continuous abstinence rate among the subset of the intervention group who took up the offer of the app was 80% higher than the comparator group (12.7% vs. 7.0%; **Table 3)**. After adjustment for baseline covariates (age, financial status, level of addiction, and current use of evidence-based support), the difference between groups attenuated to 60% but remained statistically significant (**Table 3**). Among this subset of the intervention group, the rate of quit attempts was 40% higher than the comparator group (21.5% vs. 15.3%), the rate of 3-month continuous abstinence was 85% higher (6.3% vs. 3.4%), and the rate of 6-month continuous abstinence among those who made a quit attempt was 60% higher (30.3% vs. 19.8%); these differences remained statistically significant after adjustment for covariates (**Table 3**).

**Table 3.**
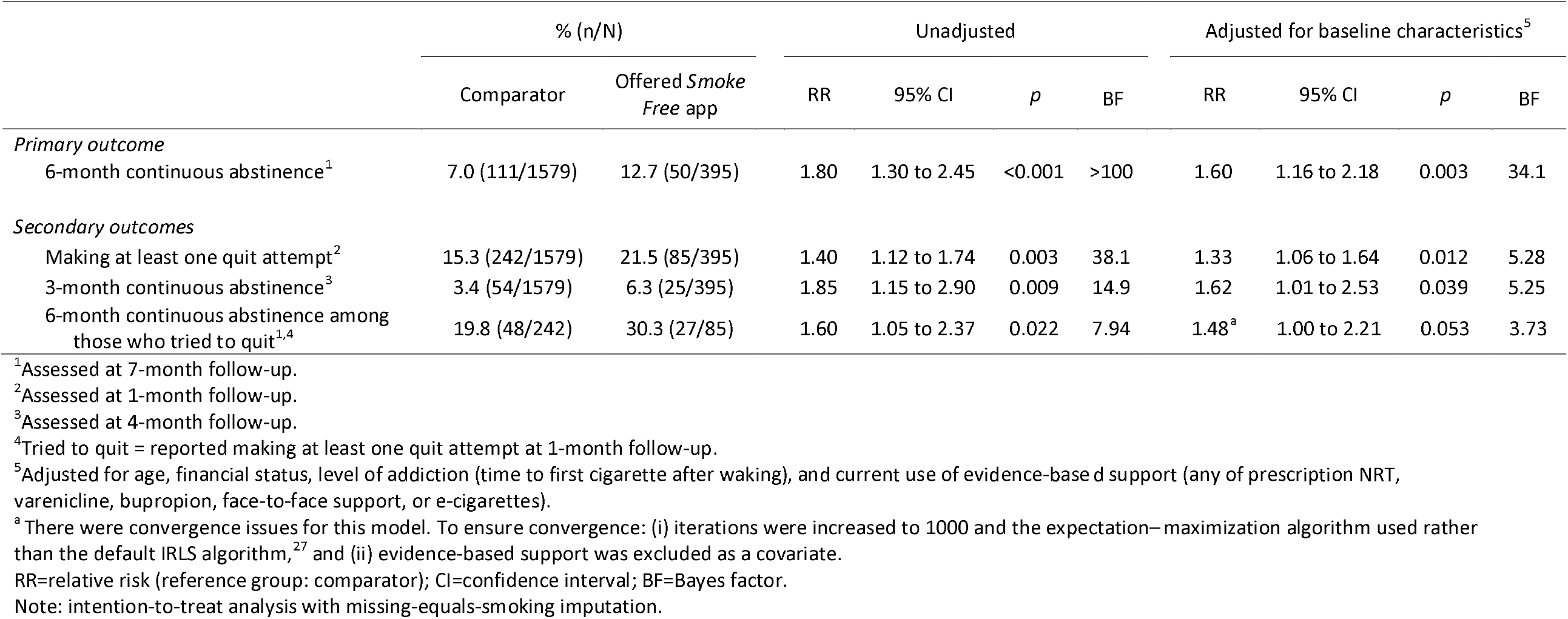
Sensitivity analysis: restricting intervention group to those who took up the offer of the app (self-reported or verified)

**Table 4** shows results for primary and secondary outcomes with analyses run on complete cases. There was no statistically significant difference between groups in rates of 6-month continuous abstinence, making at least one quit attempt, 3-month continuous abstinence, or 6-month continuous abstinence among those who tried to quit (Bayes factors for these outcomes indicated the data were insensitive or favoured no effect). Participants in the intervention group were twice as likely to report having downloaded or used the *Smoke Free* app at least once during the study period.

**Table 4.** Sensitivity analysis: complete cases

|  | % (n/N) |  | RR | 95% CI | *p* | BF |
| --- | --- | --- | --- | --- | --- | --- |
|  | Comparator | Offered *Smoke Free* app |  |  |  |  |
| *Primary outcome* |  |  |  |  |  |  |
| 6-month continuous abstinence<sup>1</sup> | 15.6 (111/712) | 17.6 (107/609) | 1.13 | 0.88 to 1.44 | 0.334 | 0.75 |
| *Secondary outcomes* |  |  |  |  |  |  |
| Making at least one quit attempt<sup>2</sup> | 68.4 (242/354) | 70.6 (180/255) | 1.03 | 0.93 to 1.15 | 0.555 | 0.22 |
| 3-month continuous abstinence<sup>3</sup> | 12.3 (54/438) | 14.6 (45/309) | 1.18 | 0.81 to 1.70 | 0.375 | 0.91 |
| 6-month continuous abstinence among those who tried to quit<sup>1,4</sup> | 21.3 (48/225) | 26.9 (43/160) | 1.26 | 0.88 to 1.80 | 0.206 | 1.41 |
| Reported downloading or using the *Smoke Free* app at least once<sup>1</sup> | 24.1 (51/212) | 48.3 (85/176) | 2.01 | 1.52 to 2.69 | <0.001 | >100 |

<sup>1</sup>Assessed at 7-month follow-up.<sup>2</sup>Assessed at 1-month follow-up.<sup>3</sup>Assessed at 4-month follow-up.<sup>4</sup>Tried to quit = reported making at least one quit attempt at 1-month follow-up.
RR=relative risk (reference group: comparator); CI=confidence interval; BF=Bayes factor.

**Table 5** shows results for primary and secondary outcomes with missing data imputed using multiple imputation. Results were very similar to the complete-case analysis: there was no statistically significant difference between groups in rates of 6-month continuous abstinence, making at least one quit attempt, 3-month continuous abstinence, or 6-month continuous abstinence among those who tried to quit (Bayes factors for these outcomes indicated the data were insensitive or favoured no effect). Participants in the intervention group were 72% more likely to report having downloaded or used the *Smoke Free* app at least once during the study period.

**Table 5.** Sensitivity analysis: multiple imputation

|  | % (n/N) |  | RR | 95% CI | *p* | BF |
| --- | --- | --- | --- | --- | --- | --- |
|  | Comparator | Offered *Smoke Free* app |  |  |  |  |
| *Primary outcome* |  |  |  |  |  |  |
| 6-month continuous abstinence<sup>1</sup> | 20.0 (316/1579) | 22.4 (350/1564) | 1.12 | 0.94 to 1.33 | 0.206 | 0.84 |
| *Secondary outcomes* |  |  |  |  |  |  |
| Making at least one quit attempt<sup>2</sup> | 63.5 (1002/1579) | 61.3 (959/1564) | 0.97 | 0.87 to 1.07 | 0.488 | 0.08 |
| 3-month continuous abstinence<sup>3</sup> | 17.0 (269/1579) | 19.1 (299/1564) | 1.12 | 0.91 to 1.39 | 0.290 | 0.72 |
| 6-month continuous abstinence among those who tried to quit<sup>1,4</sup> | 23.6 (241/1022) | 26.5 (254/959) | 1.22 | 0.85 to 1.74 | 0.277 | 1.14 |
| Reported downloading or using the Smoke Free app at least once<sup>1</sup> | 26.4 (416/1579) | 45.3 (708/1564) | 1.72 | 1.39 to 2.13 | <0.001 | >100 |

<sup>1</sup>Assessed at 7-month follow-up.<sup>2</sup>Assessed at 1-month follow-up.<sup>3</sup>Assessed at 4-month follow-up.<sup>4</sup>Tried to quit = reported making at least one quit attempt at 1-month follow-up.
RR=relative risk (reference group: comparator); CI=confidence interval; BF=Bayes factor.

A total of 108 participants reported their resting heart rate at baseline and 7-month follow-up. When we defined successful quits as self-reported abstinence plus a reduction in mean resting heart rate of ≥5 beats per minute, the rate of 6-month continuous abstinence did not differ significantly between the intervention and comparator arms (0.32% [5/1564] vs. 0.57% [9/1579]; RR=0.56, 95%CI=0.17-1.62, p=0.299). The Bayes factor indicated the data were insensitive (BF=0.51).

Assuming different rates of abstinence among participants who were lost to follow-up had little effect on our primary outcome (**Supplementary File 5**).

## Discussion

Among motivated smokers provided with very brief advice to quit, there was no significant difference in 6-month smoking cessation rates between participants randomised to receive the offer of the *Smoke Free* app plus follow-up and those randomised to follow-up only. This result was observed across analyses using intention-to-treat with missing-equals-smoking imputation, complete cases, and multiple imputation, and there were similar results on the secondary outcomes. However, when the intervention group was restricted to those who took up the offer of the *Smoke Free* app a significant benefit of treatment was observed, with participants in the intervention group 80% more likely to report abstinence than those in the comparator group on the primary outcome, with similar results on secondary outcomes. This was only partly explained by differences in baseline characteristics, with the effect remaining at 60% (a statistically significant difference) after adjustment for age, financial status, level of addiction, and current use of evidence-based cessation support.

There was no significant difference in 6-month continuous abstinence among those who tried to quit in intention-to-treat, complete case, and multiply imputed analyses. These analyses were limited by low response to the 1-month follow-up survey (which assessed quit attempts) and Bayes factors indicated the data were insensitive. The data showed a benefit of treatment when the intervention group was restricted to those who took up the offer of the *Smoke Free* app, with the intervention group 60% more likely to report abstinence than those in the comparator group.

Intention-to-treat analyses indicated a lower rate of quit attempts in the first four weeks in the intervention group compared with the comparator. However, this was not consistently observed across sensitivity analyses that used complete cases or multiple imputation, which showed no significant effect (with Bayes factors favouring no difference). It is likely the intention-to-treat result was an artefact resulting from the lower response rate to the 1-month follow-up survey in the intervention versus comparator group (16% vs. 23%, meaning a greater proportion of the intervention group were assumed not to have made a quit attempt) rather than any genuine difference between the groups. Indeed, when the intervention group was restricted to those who took up the offer of the *Smoke Free* app, a significant benefit of treatment was observed, with participants in the intervention group 40% more likely to report attempting to quit.

There was no statistically significant difference in 3-month continuous abstinence rates between groups. Analyses of this outcome were limited by the low response rate to the 4-month follow-up survey (16%). Bayes factors indicated the data were insensitive for all analyses except the intention-to-treat analysis, which favoured no effect, and the analysis restricting the intervention group to those who took up the offer of the app, which showed a significant benefit of treatment (85% more likely to report 3-month continuous abstinence).

Participants in the intervention group were significantly more likely to report having downloaded or used the *Smoke Free* app at least once during the study period, although uptake of the offer of the app was low across self-report (5%) and validated (23%) measures (25% overall). Prevalence of self-reported uptake was suppressed by the low response to the final follow-up survey, particularly because many responders (i.e., those who responded via email/telephone) only provided data on the primary outcome. However, the validated measure of treatment uptake will have captured the majority participants who took up the offer of the app, as long as they used the same email address to sign up to the study and register for the app.

To our knowledge, this is the first study to test the effectiveness of a smoking cessation app compared with unaided quitting. It differs from other large trials of smoking cessation apps^11^ not only in its comparator group (i.e., follow-up only rather than active treatment) but also in the way it was advertised, making no reference to smartphone apps until participants were enrolled. It did not aim to target smokers interested in quitting with the support of an app, but rather any smoker motivated to make a quit attempt. Thus, the relatively low uptake of the offer of the app in the intervention group is not surprising. Representative observational data show low rates of adoption of digital aids for smoking cessation, with fewer than 3% of smokers who have tried to quit reporting using a digital cessation aid (app or website).^28^ Our results suggest that while not every smoker will be interested in trying them, use of smoking cessation apps can be increased by directing smokers to this type of support (25% of those offered registered an account with the app). Our data also show that the *Smoke Free* app boosted quit rates among smokers who used it. Given the wide reach of smartphone apps, it is possible that initiatives to increase smokers’ awareness of smoking cessation apps could have a meaningful impact on rates of cessation at the population level even if only a minority of smokers take up use of app-based support (even a small percentage of a very large number can be a large number). Analyses based on complete cases and multiply imputed data indicated offering the app increased risk of 6-month continuous abstinence by 10% versus follow-up only. While this would generally be considered a small effect size for a behavioural intervention, small effects of treatments that aid smoking cessation can be clinically significant because of the very large health gains that accrue from stopping smoking.^29^ Moreover, offering the app to all smokers at a population level (a low-cost, highly scalable intervention) could result in a large number of successful quits.

Strengths of this study include the large sample size, the wide geographic scope and the 6-month follow-up duration.^22^ The pragmatic design offers real-world insights, focusing on the offer rather than use of the app, as not every smoker will want to use digital support or apps. There were also several limitations. First, there was a high rate of attrition. As the study adopted a pragmatic design and involved a very light touch intervention, we anticipated that response to follow-up attempts would be relatively low^16^ and concentrated our limited resources on maximising response to the final follow-up at 7 months. Secondly, our study should have been powered for a smaller effect size given the low rate of uptake in the intervention group. With just 25% of participants taking up the offer of the app and an observed effect size of RR∼1.3-1.5 among this group, future trials of this nature would need to be powered to detect smaller effects (RR∼1.1) which would require samples in the region of 64,000 participants. This is not unfeasible given the numbers involved in some digital trials.^30^ Future studies could investigate barriers to app use to explore the low rate of uptake of the offer of free access to a paid app in a motivated group of smokers. Thirdly, we did not undertake remote biochemical data collection to verify abstinence. While biochemical verification is widely considered the gold-standard for evaluating cessation outcomes,^22,31^ the Society for Research on Nicotine and Tobacco Subcommittee on Biochemical Verification has advised that in large-scale population-based trials such as this one, where face-to-face contact is limited and data are optimally collected online, the added precision gained by biochemical verification may be offset by methodological problems in such a way that its use is not required and may not be desirable.^31,32^ Given our study’s large, geographically dispersed participant sample, collecting biological samples (e.g. saliva, urine, or blood) or conducting remote observation of rapid tests would have been costly and logistically challenging. It may also have reduced the representativeness of the sample (if smokers unwilling to provide biological samples were ineligible to participate) or increased the rate of missing outcome data (if logistical complexity and participant burden reduced the likelihood of response to follow-up or resulted in unusable samples).^33^ We explored the possibility of verifying abstinence via a reduction in self-reported resting heart rate (obtained via a monitoring device such as a Fitbit), but only a small minority of participants provided this data and the analysis was insensitive. Collecting this data was not an important aspect of our follow-up strategy and it may be feasible in future trials with additional incentive. Finally, while we recruited an international sample, with no restrictions on location, the majority (97%) of participants were from five high-income Western countries (the UK, the USA, Canada, Ireland, and Australia). This may limit the generalisability of the results. Further investigation is required in low- and middle-income countries, where the potential benefits of smoking cessation apps may be greater in the absence of comprehensive and affordable cessation support. In conclusion, among motivated smokers provided with very brief advice to quit, the *Smoke Free* app did not have a detectable benefit for cessation compared with follow-up only. However, there was evidence that the app increased quit rates when smokers randomised to receive the app downloaded it.

## Data Availability

All data produced in the present study are available upon reasonable request to the authors

## Acknowledgments

We gratefully acknowledge David Crane, creator of the *Smoke Free* app, for making this research possible. We also thank our Trial Steering Committee for their thoughtful guidance throughout the duration of the project.

## Notes

### Competing Interest Statement

EB and JB have received unrestricted research funding from Pfizer, and JB only from J&J, who manufacture smoking cessation medications. RW has undertaken research and consultancy for and receives travel funds and hospitality from manufacturers of smoking cessation medications (Pfizer, GlaxoSmithKline and Johnson and Johnson). OP, RW, and JB are unpaid members of the scientific steering group of the Smoke Free mobile application. All authors declare no financial links with tobacco companies, e-cigarette manufacturers, or their representatives.

### Clinical Trial

ISRCTN85785540

### Clinical Protocols

https://onlinelibrary.wiley.com/doi/full/10.1111/add.14652

### Funding Statement

This study was funded by Cancer Research UK (PRCRPG-Nov21\100002).

### Author Declarations

Ethical approval for the trial was obtained from the UCL Research Ethics Committee (reference: CEHP/2020/579).

