## Supplementary File 3 for "Effectiveness of the offer of the *Smoke Free* smartphone application compared with no intervention for smoking cessation: a pragmatic randomised controlled trial"

**Supplementary File 3: results of moderation analyses**

**Table 1** summarises tests of moderation of treatment effects by baseline characteristics (gender, level of addiction, age, education, financial situation, and previous experience with a smoking cessation app). For the primary outcome of 6-month continuous abstinence, the treatment effect did not differ significantly according to any characteristic. However, there was some evidence of moderation of secondary outcomes.

For making a quit attempt, there was a significant interaction between group and level of cigarette addiction (**Table 1**; **Figure 1**). Stratified analyses indicated that the likelihood of making a quit attempt was only significantly lower in the *Smoke Free* group versus comparator group among participants who were more addicted (i.e. those who reported smoking their first cigarette of the day within 5 minutes of waking: RR 0.58, 95% CI 0.42-0.78, *p*<0.001, or 6-30 minutes after waking: RR 0.73, 95% CI 0.55-0.97, *p*=0.033). There was no significant group difference in quit attempts among those who smoked their first cigarette 31-60 minutes (RR 0.92, 95% CI 0.55-1.54, *p*=0.737) or >60 minutes after waking (RR 1.31, 95% CI 0.80-2.18, *p*=0.288).

|  | Comparator | Smoke Free |
| --- | --- | --- |
| Time to first cigarette (minutes) ≤5 | 13.9% | 8% |
| Time to first cigarette (minutes) 6 to 30 | 15.4% | 11.3% |
| Time to first cigarette (minutes) 31 to 60 | 18.7% | 17.1% |
| Time to first cigarette (minutes) >60 | 21.1% | 27.5% |

**Figure 1.** Rates of quit attempts by level of addiction (indexed by time to first cigarette after waking) and group

**Table 1.** Tests of moderation of treatment effect by key baseline characteristics

|  | Group*gender | | |  | Group*level of addiction | | |  | Group*age | | |
| --- | --- | --- | --- | --- | --- | --- | --- | --- | --- | --- | --- |
|  | RR | 95% CI | *p* |  | RR | 95% CI | *p* |  | RR | 95% CI | *p* |
| *Primary outcome* |  |  |  |  |  |  |  |  |  |  |  |
| 6-month continuous abstinence^1^ | 1.03 | 0.57 to 1.86 | 0.910 |  | 0.80 | 0.62 to 1.03 | 0.090 |  | 1.21 | 0.59 to 2.49 | 0.603 |
| *Secondary outcomes* |  |  |  |  |  |  |  |  |  |  |  |
| Making at least one quit attempt^2^ | 0.84 | 0.57 to 1.23 | 0.369 |  | **1.31** | **1.10 to 1.57** | **0.003** |  | 0.64 | 0.41 to 1.01 | 0.054 |
| 3-month continuous abstinence^3^ | 1.18 | 0.52 to 2.70 | 0.692 |  | 0.89 | 0.60 to 1.30 | 0.537 |  | 0.81 | 0.31 to 2.13 | 0.665 |
| 6-month continuous abstinence among those who tried to quit^1,4^ | 1.11 | 0.50 to 2.45 | 0.803 |  | 0.87 | 0.63 to 1.22 | 0.420 |  | 1.21 | 0.49 to 2.93 | 0.678 |
| Reported downloading or using the Smoke Free app at least once^1^ | 1.07 | 0.50 to 2.24 | 0.861 |  | 0.96 | 0.69 to 1.34 | 0.809 |  | **2.37** | **1.09 to 5.22** | **0.029** |
|  | Group*education | | |  | Group*financial situation | | |  | Group*previous experience with a smoking cessation app | | |
|  | RR | 95% CI | *p* |  | RR | 95% CI | *p* |  | RR | 95% CI | *p* |
| *Primary outcome* |  |  |  |  |  |  |  |  |  |  |  |
| 6-month continuous abstinence^1^ | 0.47 | 0.18 to 1.16 | 0.112 |  | 1.02 | 0.75 to 1.40 | 0.894 |  | 0.74 | 0.44 to 1.23 | 0.246 |
| *Secondary outcomes* |  |  |  |  |  |  |  |  |  |  |  |
| Making at least one quit attempt^2^ | 1.63 | 0.83 to 3.44 | 0.170 |  | 0.93 | 0.75 to 1.16 | 0.536 |  | 0.85 | 0.94 to 1.50 | 0.369 |
| 3-month continuous abstinence^3^ | 0.80 | 0.14 to 4.43 | 0.784 |  | 0.97 | 0.59 to 1.58 | 0.890 |  | 0.76 | 0.35 to 1.66 | 0.489 |
| 6-month continuous abstinence among those who tried to quit^1,4^ | **0.20** | **0.03 to 0.79** | **0.036** |  | 1.24 | 0.80 to 1.92 | 0.337 |  | 0.61 | 0.29 to 1.28 | 0.195 |
| Reported downloading or using the Smoke Free app at least once^1^ | 1.22 | 0.24 to 5.67 | 0.795 |  | 0.90 | 0.59 to 1.36 | 0.607 |  | 1.07 | 0.50 to 2.24 | 0.861 |

^1^Assessed at 7-month follow-up.

^2^Assessed at 1-month follow-up.

^3^Assessed at 4-month follow-up.

^4^Tried to quit = reported making at least one quit attempt at 1-month follow-up.

Note: intention-to-treat analysis with missing-equals-smoking imputation. Independent variables were coded as follows: group (0=comparator, 1=Smoke Free), gender (0=male, 1=female), level of addiction (time to first cigarette: 0=within 5 minutes, 1=6-30 minutes, 2=31-60 minutes, 4=after 60 minutes), age (0=<35, 1=≥35), education (0=no post-16 qualifications, 1=post-16 qualifications), financial situation (0=don’t meet basic expenses, 1=just meet basic expenses, 2=meets needs with a little left, 3=live comfortably), previous experience with a smoking cessation app (0=no, 1=yes). Bold font indicates statistically significant interactions.

For 6-month continuous abstinence among those who tried to quit, there was a significant interaction between group and education (**Table 1**, **Figure 2**). While the risk of abstinence did not differ significantly by group at either level of education, the point estimate was lower among those with post-16 qualifications (RR 1.07, 95% CI 0.73-1.56, *p*=0.743) than those without (RR 5.45, 95% CI 1.37-36.73, *p*=0.036).

|  | Comparator | Smoke Free |
| --- | --- | --- |
| Post-16 qualifications No | 8.3% | 45.5% |
| Post-16 qualifications Yes | 21.1% | 22.5% |

**Figure 2.** Rates of 6-month continuous abstinence among those who tried to quit by level of education and group

For downloading or using the *Smoke Free* app at least once during the study period, there was a significant interaction between group and age (**Table 1**; **Figure 3**). The risk of app use was significantly higher in the *Smoke Free* group versus comparator group among older participants (≥35 years: RR 2.08, 95% CI 1.40-3.15, *p*<0.001) but did not differ significantly by group among those aged <35 (RR 0.88, 95% CI 0.45-1.71, *p*=0.699).

|  | Comparator | Smoke Free |
| --- | --- | --- |
| Age <35 | 9.4% | 8.3% |
| Age ≥35 | 2.4% | 5.1% |

**Figure 3.** Rates of reporting downloading or using the Smoke Free app at least once by age and group
