## Supplementary File 4 for "Effectiveness of the offer of the *Smoke Free* smartphone application compared with no intervention for smoking cessation: a pragmatic randomised controlled trial"

**Supplementary File 4: baseline characteristics, restricting intervention group to those who took up the offer of the app**

**Table 1.** Baseline characteristics of participants randomised to each condition restricting the intervention group to those who took up the offer of the app

|  |  | Comparator (n=1579) | |  | Took up the offer of the *Smoke Free* app (n=395) | |
| --- | --- | --- | --- | --- | --- | --- |
|  |  | % | n |  | % | N |
| Age (years) |  |  |  |  |  |  |
| 18-34 |  | 11.4 | 180 |  | 15.4 | 61 |
| 35-64 |  | 82.0 | 1294 |  | 79.2 | 313 |
| 65+ |  | 6.7 | 105 |  | 5.3 | 21 |
| Gender |  |  |  |  |  |  |
| Male |  | 24.2 | 382 |  | 28.9 | 114 |
| Female |  | 75.5 | 1192 |  | 70.6 | 279 |
| Other |  | 0.3 | 5 |  | 0.2 | 2 |
| Post-16 qualifications |  | 90.4 | 1427 |  | 95.2 | 376 |
| Financial status |  |  |  |  |  |  |
| Live comfortably |  | 5.9 | 93 |  | 5.6 | 2 |
| Meet needs with a little left |  | 33.5 | 529 |  | 29.6 | 117 |
| Just meet basic expenses |  | 39.7 | 627 |  | 43.3 | 171 |
| Don’t meet basic expenses |  | 20.9 | 330 |  | 21.5 | 85 |
| Country of residence |  |  |  |  |  |  |
| UK |  | 45.8 | 723 |  | 51.1 | 202 |
| USA |  | 34.3 | 542 |  | 26.3 | 104 |
| Canada |  | 8.0 | 127 |  | 8.6 | 34 |
| Ireland |  | 5.8 | 91 |  | 6.8 | 27 |
| Australia |  | 3.0 | 48 |  | 3.8 | 15 |
| Other |  | 3.0 | 48 |  | 3.3 | 13 |
| English as first language |  | 95.8 | 1513 |  | 92.7 | 366 |
| Time to first cigarette |  |  |  |  |  |  |
| ≤5 minutes |  | 46.0 | 726 |  | 41.0 | 162 |
| 6-30 minutes |  | 40.2 | 635 |  | 37.5 | 148 |
| 31-60 minutes |  | 7.8 | 123 |  | 11.5 | 45 |
| >60 minutes |  | 6.0 | 95 |  | 10.1 | 40 |
| History of serious quit attempts |  |  |  |  |  |  |
| Never |  | 7.3 | 115 |  | 6.6 | 26 |
| Yes – not in the past year |  | 59.6 | 941 |  | 56.5 | 223 |
| Yes – in the past year |  | 33.1 | 523 |  | 37.0 | 146 |

*Table continued on next page.*

**Table 1.** (continued)

|  |  | Comparator (n=1579) | |  | Took up the offer of the *Smoke Free* app (n=355) | |
| --- | --- | --- | --- | --- | --- | --- |
|  |  | % | n |  | % | n |
| Past use of cessation support |  |  |  |  |  |  |
| Prescription NRT |  | 53.1 | 839 |  | 53.7 | 212 |
| NRT bought over the counter |  | 30.0 | 474 |  | 29.6 | 117 |
| Varenicline |  | 15.6 | 246 |  | 21.3 | 84 |
| Bupropion |  | 14.0 | 222 |  | 16.5 | 65 |
| Face-to-face behavioural support |  | 8.7 | 138 |  | 9.6 | 38 |
| Telephone support |  | 5.9 | 93 |  | 8.6 | 34 |
| Written self-help materials |  | 24.3 | 384 |  | 24.6 | 97 |
| Websites |  | 11.0 | 173 |  | 12.2 | 48 |
| Apps |  | 47.7 | 753 |  | 55.2 | 218 |
| E-cigarette or other vaping device |  | 17.0 | 269 |  | 24.1 | 95 |
| Other |  | 2.7 | 43 |  | 4.1 | 16 |
| None of the above |  | 13.1 | 206 |  | 12.7 | 50 |
| Current use of cessation support |  |  |  |  |  |  |
| Prescription NRT |  | 2.5 | 39 |  | 3.0 | 12 |
| NRT bought over the counter |  | 9.1 | 143 |  | 8.6 | 34 |
| Varenicline |  | 2.0 | 31 |  | 3.0 | 12 |
| Bupropion |  | 0.7 | 11 |  | 0.8 | 3 |
| Face-to-face behavioural support |  | 0.1 | 2 |  | 0.3 | 1 |
| Telephone support |  | 0.4 | 7 |  | 1.3 | 5 |
| Written self-help materials |  | 1.8 | 28 |  | 1.5 | 6 |
| Websites |  | 2.4 | 38 |  | 3.8 | 15 |
| Apps |  | 4.0 | 63 |  | 6.3 | 25 |
| E-cigarette or other vaping device |  | 12.3 | 194 |  | 13.4 | 53 |
| Other |  | 0.8 | 12 |  | 1.0 | 1 |
| None of the above |  | 70.6 | 1114 |  | 65.3 | 258 |
|  |  | Mean | SD |  | Mean | SD |
| Age (years) |  | 48.9 | 11.4 |  | 47.3 | 11.5 |
| Cigarettes per day |  | 18.2 | 10.0 |  | 17.4 | 8.5 |
| Resting heart rate* |  | 75.4 | 18.7 |  | 75.0 | 18.0 |

NRT, nicotine replacement therapy. SD, standard deviation.

* If participants had a heart monitoring device (e.g. Fitbit, Apple watch); this was not a required field
