## Supplementary File 5 for "Effectiveness of the offer of the *Smoke Free* smartphone application compared with no intervention for smoking cessation: a pragmatic randomised controlled trial"

**Supplementary File 5: impact of assuming different rates of abstinence in participants lost to follow-up**

**Table 1.** Sensitivity analysis testing the impact of assuming different rates of abstinence in those not followed-up on the primary outcome

| **Imputed abstinence rate in missing*** | **Group** | **Missing / N** | **Abstinence rate (n)†** | **RR† [95% CI]** |
| --- | --- | --- | --- | --- |
| 0% | Comparator | 870 / 1579 | 7.03% (111.0) | Ref |
|  | *Smoke Free* | 957 / 1564 | 6.84% (107.0) | 0.97 [0.75 to 1.13] |
| 10% | Comparator | 870 / 1579 | 12.5% (198.0) | Ref |
|  | *Smoke Free* | 957 / 1564 | 13.0% (202.7) | 1.04 [0.86 to 1.24] |
| 20% | Comparator | 870 / 1579 | 18.0% (285.0) | Ref |
|  | *Smoke Free* | 957 / 1564 | 19.1% (298.4) | 1.06 [0.91 to 1.22] |
| 30% | Comparator | 870 / 1579 | 23.6% (372.0) | Ref |
|  | *Smoke Free* | 957 / 1564 | 25.2% (394.1) | 1.07 [0.95 to 1.21] |
| 40% | Comparator | 870 / 1579 | 29.1% (459.0) | Ref |
|  | *Smoke Free* | 957 / 1564 | 31.3% (489.8) | 1.08 [0.97 to 1.20] |

*	Imputed abstinence rate among participants who were missing at 7-month follow-up and did not report having returned to smoking at 1- or 4-month follow-up.

†	Estimated percentage and number (n) of people abstinent from cigarette smoking for 6 months, after imputing the abstinence rate in those missing at follow-up. Risk ratio (RR) calculated from these estimates.
