## Supplementary File 1 for "Effectiveness of the offer of the *Smoke Free* smartphone application compared with no intervention for smoking cessation: a pragmatic randomised controlled trial"

**Supplementary File 1: additional information on methods**

**Contents:**

- Study timeline
- Recruitment advert posted on Facebook and Twitter
- Message shown to both intervention and comparator groups at the end of the baseline survey

**Study timeline**

| ***Month*** | **0** | **1** |  |  | **4** |  |  | **7** |
| --- | --- | --- | --- | --- | --- | --- | --- | --- |
|  | - Recruitment - Consent - Screening - Baseline questionnaire - Randomisation - Brief message encouraging making a quit attempt | - 1-month follow-up questionnaire |  |  | - 4-month follow-up questionnaire |  |  | - 7-month follow-up questionnaire |

**Recruitment advert posted on Facebook and Twitter**

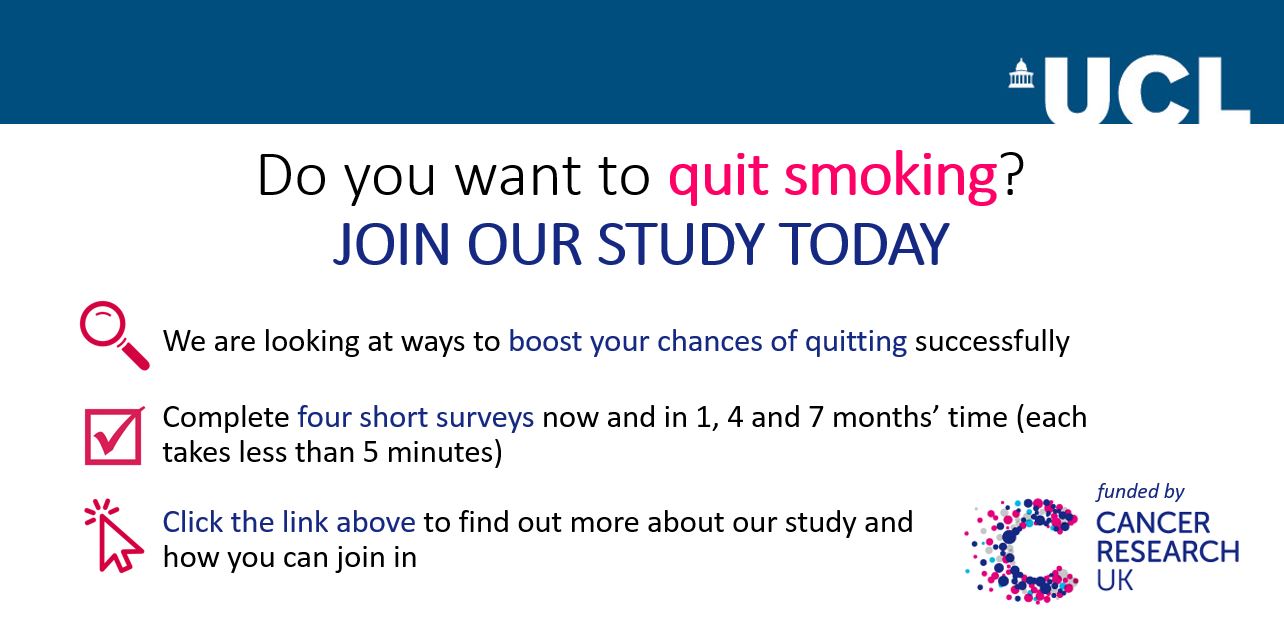

**Message shown to both intervention and comparator groups at the end of the baseline survey.**

You said you wanted to make a quit attempt in the next 4 weeks. That’s great! Quitting is the single best thing you can do for your health. When will you start? Think carefully about the best date and time for you. It might be right now.

The first 24 hours are crucial - get past those and you’ll be twice as likely to stay off cigarettes for good! One thing that we know helps is making a serious commitment to being a non-smoker. Keeping track of your progress is another good technique.

Thank you again for completing the survey and taking part in our study. We’ll send you an email in a few weeks to ask how you’re getting on. Please reply, it makes a big difference to our Cancer Research UK funded study if you do.
