## Supplementary File 2 for "Effectiveness of the offer of the *Smoke Free* smartphone application compared with no intervention for smoking cessation: a pragmatic randomised controlled trial"

**Supplementary File 2: amendments to the analysis plan**

We followed our pre-registered analysis plan^1^, with two amendments registered on Open Science Framework prior to running the analysis (<https://osf.io/umec4/>):

1. We added another sensitivity analysis. On the suggestion of our Trial Steering Committee, we used multiple imputation of missing outcome data.
2. We amended one of our moderation analyses. We had intended to test for moderation by previous experience with the *Smoke Free* app by matching the email address provided by participants at baseline with the mailing list of previous *Smoke Free* users we used for recruitment. Because we recruited a smaller proportion of participants than expected from the mailing list of previous *Smoke Free* users, meaning previous exposure to this app specifically was likely to be less prevalent, we amended this analysis to look at moderation by previous experience with any smoking cessation app.

We also made two further amendments after running the analyses:

3. We compared the baseline characteristics of the participants in the intervention group who took up the offer of the app with those of the comparator group, and repeated the sensitivity analysis restricting the intervention group to those who took up the offer of the app with the addition of adjustment for baseline characteristics. Because we assessed a large number of variables at baseline, we limited the adjustment to variables known to be associated with success in quitting smoking:^2,3^ age, financial status (a marker of socioeconomic position), time to first cigarette after waking (a marker of level of cigarette addiction), and current use of evidence-based support (coded 1 for any of prescription NRT, varenicline, bupropion, face-to-face support, or e-cigarettes and 0 for none of these).
4. We ran an additional sensitivity analysis in which we explored the impact of assuming different rates of abstinence in those not followed-up on our primary outcome.
